# Mental Illness Self-Stigma in Young People: A Scoping Review Protocol

**DOI:** 10.1101/2021.07.06.21260070

**Authors:** Rachel Taylor, Nicola Cogan, Pamela Jenkins, Xi Liu, Paul Flowers, Simon C. Hunter, Patrick Corrigan

**Affiliations:** University of Strathclyde; Mental Health Foundation Scotland; Glasgow Caledonian University; Illinois Institute of Technology

**Keywords:** Self-stigma, Mental Illness, Mental Health, Help-seeking, Young People, Scoping Review

## Abstract

**Background:** Self-stigma (the internalisation of negative stereotypes) is known to reduce help-seeking behaviours and treatment adherence in people who have a mental illness, resulting in worsening health outcomes. Moreover, self-stigma diminishes self-esteem and self-efficacy, and leads to higher levels of depression. Half of all lifetime cases of mental illness have manifested by the age of 14, and therefore young people are vulnerable to the negative impact of suffering mental illness self-stigma. While literature in this field has been flourishing in recent years, mental illness self-stigma remains poorly understood in youth. It is important that we seek to understand what is currently known about mental illness self-stigma in young people, and subsequently use this information to guide future research to advance knowledge of this topic. To date, a scoping review which maps the available literature on mental illness self-stigma in young people has yet to be conducted. Here, we outline the protocol for a scoping review on mental illness self-stigma in young people.

**Methods:** This protocol outlines the process of conducting a scoping review of primary research concerning mental illness self-stigma in young people. The Joanna Briggs Institute guidance on best practice for conducting a scoping review will utilised throughout. A systematic search of appropriate databases will be conducted which will allow for a two-step screening process and data charting. Inclusion criteria for this review dictates that eligible studies will i) include a population within the age range of 10-35 (or mean age within this range), ii) include the term ‘self-stigma’ or ‘internalised stigma’ relating to mental health/illness/disorder, iii) be primary research, iv) be published post-2009 (following the seminal study by Moses, 2009) and v) be published in English.

**Discussion:** The findings of the scoping review outlined in this protocol will be used to inform future research which aims to explore self-stigmatising attitudes and beliefs of young people experiencing mental illness. This research will result in co-produced, impactful resources for young people on the topic of mental illness self-stigma with the aim of raising awareness and stigma reduction.

---

Stigma surrounding the topic of mental illness is a significant barrier to seeking help and treatment (Pescosolido & Boyer, 1999; Pescosolido, Gardner, & Lubell, 1998, cited in Moses, 2009). Adolescence represents a critical period of identity formation, where early intervention for developing mental illness is of vital importance (Meiser & Esser, 2017, cited in Berry & Greenwood, 2018). Experiencing stigma at this age may be particularly damaging and result in worsening health outcomes (Yang et al., 2010). The Mental Health of Children and Young People (MHCYP, 2020) survey in England found that one-in-six children (aged 5-16 years) and one-in-five young people (aged 17-22 years) had a ‘probable mental disorder’. Whilst the COVID-19 pandemic is not a central focus of the current review, it is important to recognise the climate in which this research is being conducted. YoungMinds (2021) found that 67% of young people with ‘mental health needs’ believed that the COVID-19 pandemic will have a long-term negative effect on their mental health. This indicates that the mental health of young people is of increasing concern. When considering mental illness, the role of stigma is central to understanding individuals’ experiences, attitudes, and self-beliefs.

Goffman (1963), a seminal author in the field, recognised the importance of defining the concept of stigma. He wrote that stigma arises when an individual possesses an attribute which is viewed as undesirable, reducing our opinion of that individual from being a whole person to being defined by their undesirable characteristic. Discrediting an individual because of one attribute can be harmful in several ways, particularly when an individual is stigmatised against because of their mental illness. Mental illness stigma can negatively impact a person’s life; including their independence, aspirations, career, self-concept, and overall health (Arboleda-Flo’rez & Sartorius, 2008; Hinshaw, 2007, cited in Corrigan et al., 2016). Stigma manifests itself in various forms, however this scoping review will focus on self-stigma relating to mental illness. Self-stigma, or *internalised stigma*, refers to the internalisation of negative stereotypes expressed through public stigma towards individuals with ‘devalued characteristics’, such as those who suffer with mental illness (Corrigan et al., 2009). Corrigan and Watson (2002) highlight that public stigma versus self-stigma are conceptually distinct concepts. Public stigma can manifest itself in the form of social rejection or discrimination and self-stigma results from the internalisation of these experiences and the application of those beliefs to oneself. Self-stigma is thought to arise through a three-step formation process whereby individuals become aware of negative stereotypes, agree with these beliefs, and subsequently apply them to themselves (Corrigan et al., 2006). This process is known to result in diminished self-esteem and feelings of hopelessness (Corrigan et al., 2012, 2016). Tucker (2013) conducted a confirmatory factor analysis which suggested that mental illness self-stigma and self-stigma relating to help-seeking are distinct concepts. However, commonly used measures of mental illness self-stigma, such as Self-Stigma Scale (Moses, 2009), incorporate items relating help-seeking as well as mental illness stigma. Subsequently, when reviewing literature on this topic, it is difficult to disentangle the constructs of mental illness self-stigma versus stigma surrounding seeking psychological help. For the purpose of this scoping review, literature which also discusses help-seeking will be included. The scoping review will explore how current literature conceptualises mental illness self-stigma and how this manifests in a young population.

Stigma, in particular self-stigma, can prevent help-seeking, lead to poorer treatment adherence and can result in worsening health outcomes (Holubova et al., 2016a, 2016b; Ng et al., 2008; Sedlácková et al., 2015, cited in Abo-Rass et al., 2020). Individuals who experience mental illness self-stigma have reported viewing the process of seeking psychological help as a threat to their sense of self-worth and overall confidence (Kaya et al., 2015; Vogel & Wade, 2009, cited in Eagle et al., 2020). Research has identified that *fear* of stigma, specifically surrounding mental illness labels, is a significant barrier which prevents help-seeking, resulting in unmet health needs (Yeh et al., 2003), and high levels of mental illness self-stigma is associated with non-use of medication and poorer medication adherence (Fadipe et al., 2020). Participating in treatment can exacerbate levels of self-self-stigma and result in the “why try” effect (Corrigan et al., 2014). The “why try” effect is illustrative of the harm that internalising stigmatizing beliefs can have. Self-stigma is known to decrease a person’s confidence in their own abilities leading to diminished self-esteem and self-efficacy. This results in a sense of behavioural futility where individuals begin to question whether it is worth pursuing their goals i.e., “why try to get a job? I’m not worthy” (Corrigan et al., 2016). Similarly, the “why try” effect can result in questioning the validity and purpose of participating in treatment and an overall belief that treatment will not be successful (Corrigan et al., 2014). Experiencing the “why try” effect is associated with higher levels of depression, diminished capabilities, and reduced rate of recovery from mental illness (Corrigan et al., 2016; Corrigan et al., 2019). Understanding the “why try” effect has offered insight into adults’ resistance to seek treatment or help when suffering from mental illnesses and associated stigma. However, the “why try” effect has not been fully explored in a young population. Overall, literature on mental illness self-stigma overwhelmingly indicates harmful effects but rarely focuses specifically on young people. This scoping review aims to map the available evidence on this topic and highlight areas which need to be explored in future study, including the impact of mental illness self-stigma.

It is particularly important to focus on mental illness self-stigma within a young population because half of all lifetime cases of mental illness have developed by the age of 14 (Sawyer et al., 2000; Kessler et al., 2005a, 2005b, cited in Kaushik et al., 2016). Similarly, a large-scale meta-analysis found that the peak age for onset of mental illness was 14.5 years (Solmi et al., 2021). The UNFPA defines adolescents as being aged between 10-19 years, the African Youth Charter considers youth to be aged 15-35 and UNICEF/WHO/UNFPA state that ‘young people’ fall between the ages of 10 and 24 (United Nations, 2013). This scoping review hopes to include the broadest range of articles and therefore will define ‘young people’ as aged 10-35 years, to ensure all relevant literature can be included. This developmental stage in a young persons’ life represents a critical period of identify formation and consolidation with key support systems shifting from family to peers (Moses, 2009). It is possible that becoming aware of negative stereotypes or public stigma surrounding mental illness could be particularly damaging for people within this age range. Despite this possibility, research surrounding young peoples’ experiences of mental illness self-stigma did not emerge until 2009. Moses (2009) conducted the seminal study on self-stigma in an adolescent population and found that stigma was commonly felt by adolescents receiving treatment for their mental illness. When measuring self-stigma specifically, Moses found young people experiencing self-stigma were fearful about whether others would dislike them if they knew that they were receiving mental illness treatment. Furthermore, higher levels of self-stigma are associated with higher levels of depression and diminished self-esteem (Moses, 2009). Yang et al. (2010) suggests adolescents have an unclear self-concept and thus experiencing stigma during this period of development is particularly impactful. Moses (2009) highlighted the need for exploration of this topic and the production of appropriate interventions and resources for young people tackling self-stigma. For these reasons, understanding the prevalence and impact of mental illness self-stigma in young people is of particular importance for researchers, with an appetite for knowledge and intervention being highlighted for several years.

Prior to conducting this scoping review, literature suggests that developing a mental illness as a young age can lead to experiencing mental illness self-stigma. Self-stigma is known to result in avoidance of help-seeking or poor treatment adherence. This may be impacted by diminished self-efficacy as hypothesised by current understanding of the ‘why try’ effect, however the impact is worsening health outcomes. Overall, this scoping review will explore what is currently known about mental illness self-stigma in young people.

**Figure 1.**
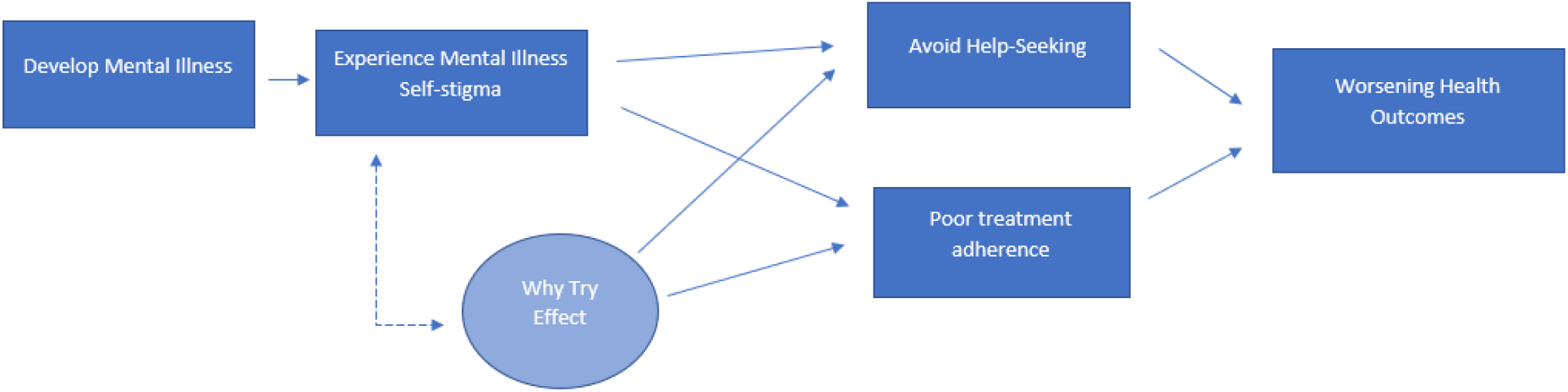

Tricco et al. (2016) reports the three most common reasons for conducting a scoping review: to explore the breadth of the literature, to summarise evidence and to inform future research. From reviewing this rationale, a scoping review is an appropriate methodology for the purpose of this research, with a key aim of mapping available literature on the topic of mental illness self-stigma in young people. By achieving this aim, it will identify novel methods for examining mental illness self-stigma in young people and facilitate the development of future studies, broadening knowledge on this under-researched topic. Scoping reviews do not aim to answer precise clinical questions and thus do not assess methodological limitations or research bias. It is important that best practice guidelines are followed throughout the process of conducting a scoping review. A working group from the Joanna Briggs Institute collated guidance to ensure that scoping review are conducted rigorously and in a transparent, trustworthy manner (Peters et al., 2015). This protocol follows the guidelines as closely as is feasible within the scope of the current project.

A preliminary search for existing scoping reviews and systematics reviews has been conducted; APA PsycInfo, Web of Science and MEDLINE were searched on 02/07/2021. As a result, 4 reviews were found; 2 systematic reviews which focused generally on stigma in young people (Kaushik et al., 2016; Ferrie et al., 2020), a systematic review protocol exploring stigma in Middle Eastern adolescents (Mohammadzeh et al., 2020) and a systematic review on the mediating effect of self-stigma and depressive symptoms on self-harm and bullying victimisation (Karanikola et al., 2018). However, no scoping reviews or systematic reviews specifically focusing on mental illness self-stigma in young people are known to exist. Kaushik et al. (2016) found that only 3 out of the 42 papers identified in the systematic review touched on self-stigma specifically in young people with mental illnesses. These 3 studies (Moses 2009a, 2009b, 2010) which discussed self-stigma shared a sample of 60 young people. Ferrie et al., (2020) note that 10 of the 27 papers included in their review discuss self-stigma. This suggests that there is a lack of research exploring mental illness self-stigma in young people. The reviews conducted by Kaushik et al (2016) and Ferrie et al., (2020) excluded papers where participants were over the age of 19 and 20 respectively, whereas the proposed scoping review aims to map a wider range of literature spanning from age 10-35 years. The scoping review outlined in this protocol will provide a novel insight into the breath of evidence on this topic and will map the relevant literature to offer a summary of current knowledge.

The objective of this scoping review is to explore the breadth of literature that is currently available concerning mental illness self-stigma in young people, and to map evidence on this topic. The scoping review will tackle several questions to ensure that a broad understanding of current knowledge is understood, such as i) how mental illness self-stigma is conceptualised, ii) what measures are most commonly used to study mental illness self-stigma in young people, iii) characteristics of participants and iv) the impact of mental illness self-stigma on young people. While research on stigma in adults has begun to recognise the importance of including the voices of those with lived experience in the research process, few to none of the studies on self-stigma in youth use participatory community-based approaches (Nieweglowski et al., 2018). This review aims to pinpoint which study designs are commonly used in the field of mental illness self-stigma in young people. This can subsequently be used to design future research studies which will meaningfully contribute to the current body of knowledge. Lastly, the scoping review hopes to highlight gaps in the literature which represent areas for necessary and insightful future research. Overall, this scoping review will inform further research which aims to deepen understanding of mental illness self-stigma in young people. An example of the future research which could be developed as a result of this scoping review would be studies exploring mental illness self-stigma utilising a participatory approach where young people with lived experience act as meaningful partners, using appropriate measures as highlighted by the literature and including participants possessing characteristics that are currently lesser understood.

## Scoping Review Questions

### What evidence is there of mental illness self-stigma in young people?

1. How is mental illness self-stigma conceptualised in the literature?
2. Do studies assess the prevalence of mental illness self-stigma in young people, and if so, how is it measured?
3. What are the characteristics of participants included in the sources of evidence identified for the primary review question?
4. What are the study designs that underpin the literature on mental illness self-stigma in young people?
5. What is the impact of mental illness self-stigma on young people?
6. What gaps are there in the literature surrounding mental illness self-stigma in young people?

## Method

The Joanna Briggs Institute guidance on how to conduct a scoping review will be adhered to throughout. In terms of reporting style, the Preferred Reporting Items for Systematic reviews and Meta-Analyses extension for Scoping Reviews checklist (PRISMA-ScR) will be used.

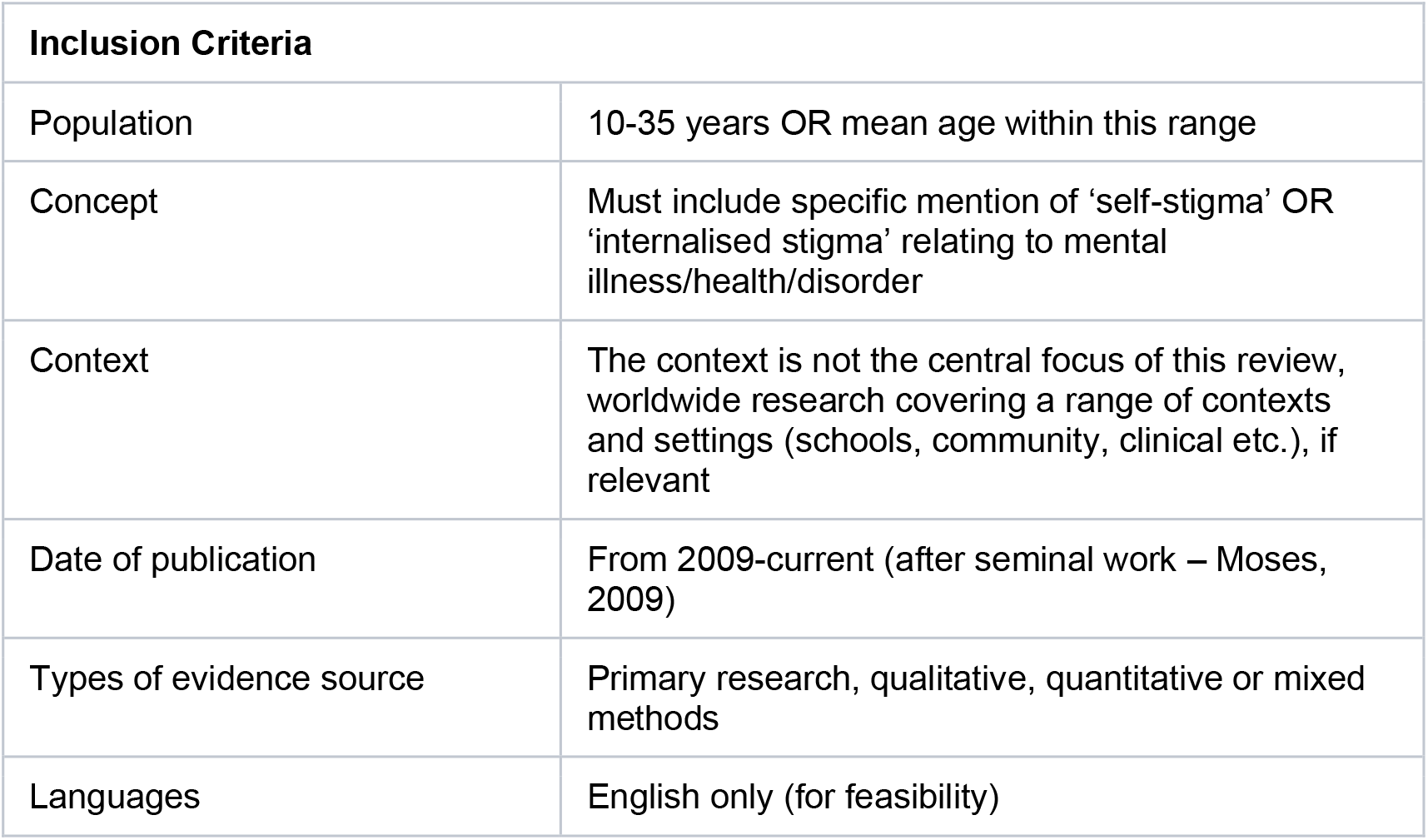

### Search Strategy

The search strategy for this scoping review will be as comprehensive as is possible and appropriate within the parameters of this project. The search terms used for this scoping review can be found in Appendix 1. Any limitations in terms of the comprehensive nature of this review will be justified in detail within the final review. In accordance with the JBI guidelines for conducting a scoping review, a three-step search strategy process will be implemented. The first step will involve performing an initial search of two databases; namely APA PsycInfo and MEDLINE (Ovid). Text words used in the title and abstracts of relevant papers identified within this search will be extracted and analysed alongside the index terms describing articles. These words/terms will then be used in step two where a second search will be conducted across all relevant databases (Cochrane library, MEDLINE (Ovid), Child and Adolescent Development Studies, Web of Science (Core Collection), APA PsycInfo). Lastly, step three will include examining reference lists of articles which have been included in the review from their full-text to identify further relevant sources. Due to the time constraints of this scoping review, authors of primary sources will not be contacted for further information and grey literature will not be explored unless it is identified during the three-step search strategy. A complete search strategy of MEDLINE will be included as an appendix of this protocol (Appendix 2).

### Sources of evidence

Two levels of screening will be used to identify sources of evidence for inclusion: 1) study selection -review title and abstract 2) study screening - review the full text. Study selection will be conducted by two independent reviewers, with the first author (RT) screening all studies and a reviewer screening 10% of studies. Any articles identified as relevant based on the title and abstract by one or both researchers will be reviewed at full-text level. The screening process involves both reviewers evaluating the full texts of selected articles in relation to the pre-determined inclusion criteria. Where disagreement occurs between reviewers, full texts will be screened again and discussion will take place until a consensus is met. If necessary, validation by a third reviewer may be sought. The software used for specifying results will be Endnote and a PRISMA flowchart will be used to report the final numbers of the study selection process,

### Data Charting

Data from articles included from full-text will be charted using a data extraction table developed by the author (see Appendix 2), based on a model recommended by the JBI. Data being charted relates to citation, country, context, participant characteristics, study aim, methodology, results, limitations, key results relating to the review questions and future areas for research. This process of data charting from the full texts will be conducted by the author (RT) and then 10% will be verified by a reviewer. Any discrepancies will be discussed until a consensus has been met, or a third reviewer may be sought.

## Results

In this scoping review, the author will extract results from included texts and map these descriptively. This method of reporting will be used to identify what is known about the key topics and to highlight any gaps in knowledge. Frequency counts for populations, concepts and characteristics will also be included. A descriptive content analysis of available literature may be conducted if appropriate based on the charted data. These methods have been chosen in accordance with the aim of this review; to explore the available information on mental illness self-stigma in young people.

### Presentation of results

Results will be presented in relation to the research questions developed for this scoping review. Accompanying these discussions, a tabular presentation of the data for aspects such as participant characteristics, study methodology, assessment tools and prevalence of mental illness self-stigma.

This scoping review will be submitted for peer-reviewed publication and will be included in a thesis for submission at the University of Strathclyde (funded by the Scottish Graduate School for Social Sciences and the Mental Health Foundation). The scoping review will inform research which will be conducted by the University of Strathclyde and the Mental Health Foundation.

## Data Availability

There is no primary data included in this submission.

## Acknowledgements

This review is undertaken as part of a PhD thesis at the University of Strathclyde under the supervision of Dr Nicola Cogan, Professor Paul Flowers, Dr Xi Liu, Professor Simon Hunter (Glasgow Caledonian University), Professor Patrick Corrigan (Illinois Institute of Technology), Dr Pamela Jenkins (Mental Health Foundation Scotland) and Lee Knifton (Mental Health Foundation).

## Appendix 1: Search Terms

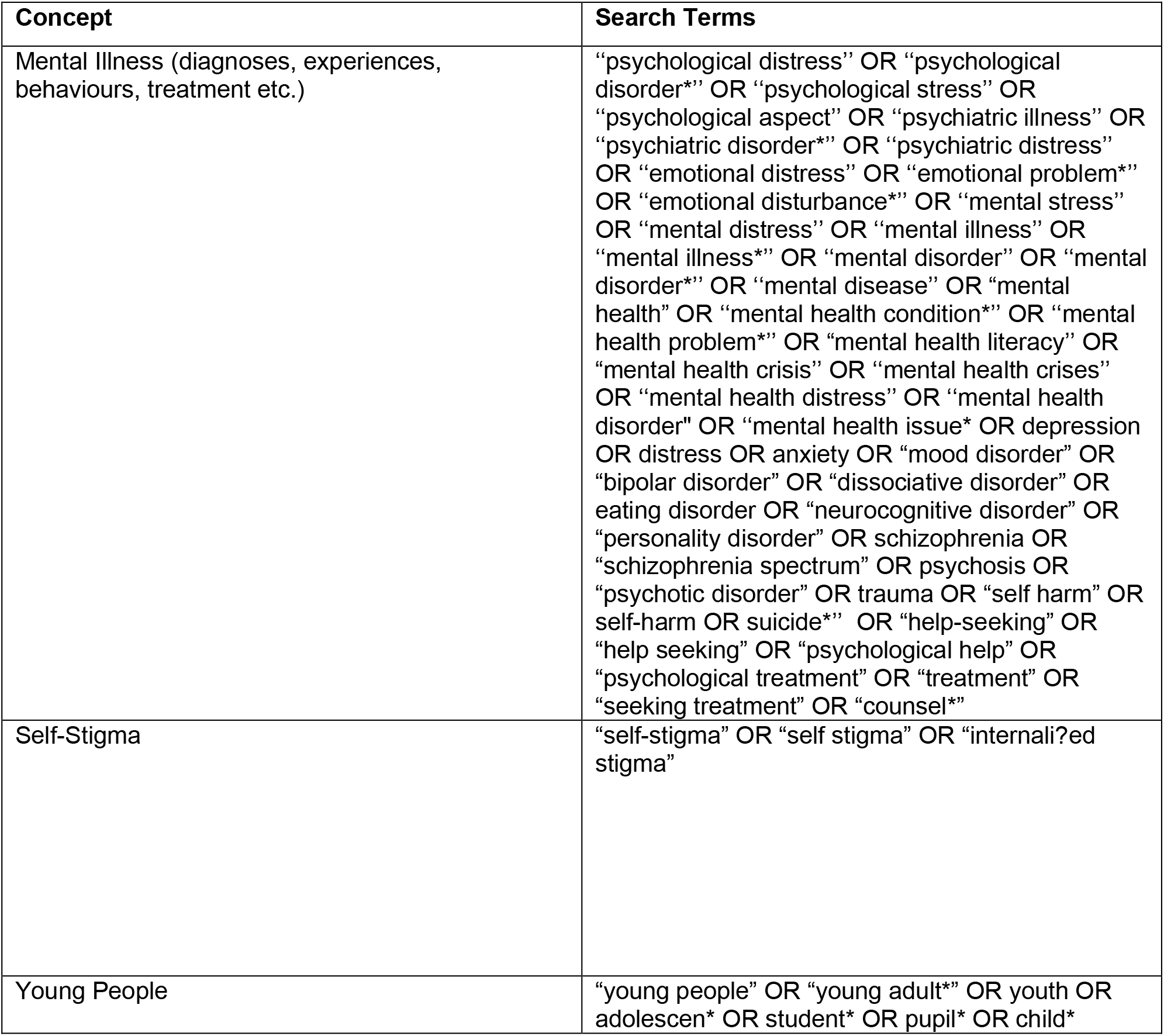

## Appendix 2: Search of MEDLINE (Ovid) – 02/07/2021

| Search | Query | Retrieved |
| --- | --- | --- |
| S1 | (psychological distress or psychological disorder* or psychological stress or psychological aspect or psychiatric illness or psychiatric disorder* or psychiatric distress or emotional distress or emotional problem* or emotional disturbance* or mental stress or mental distress or mental illness or mental illness* or mental disorder or mental disorder* or mental disease or mental health condition* or mental health problem* or mental health literacy or mental health crisis or mental health crises or mental health distress or mental health disorder or mental health issue* or depression or anxiety or mood disorder or bipolar disorder or dissociative disorder or eating disorder or neurocognitive disorder or personality disorder or schizophrenia or schizophrenia spectrum or psychosis or psychotic disorder or trauma or self harm or self-harm or suicide*).mp. | 1314436 |
| S2 | (self-stigma or self stigma or internalised stigma or internalized stigma or internali?ed stigma).mp. | 1,660 |
| S3 | (young people or young adult* or youth or adolescents or adolescen* or student* or pupil* or child* or children or childhood).mp. | 435,7836 |
| S4 | S1 AND S2 | 1130 |
| S5 | S1 AND S3 | 382775 |
| S6 | S2 AND S3 | 646 |
| S7 | S1 AND S2 AND S3 | 430 |

## Appendix 3: Extraction Table

| Evidence source Details and Characteristics |
| --- |
| Citation details (e.g. author/s, date, title, journal, volume, issue, pages) |
| Type of paper (article, review) |
| Country |
| Context (setting, location, specific interests, cultural factors) |
| Aim/Purpose of Study |
| Participant Characteristics (e.g. number, age/sex, ethnicity, sociodemographic) |
| Methodology (design, measures, duration, analysis technique) |
| Did the study use a participatory approach? (peer research, co-production etc) |
| **Details/Results extracted from source of evidence** (in relation to the research questions identified for this scoping review) |
| Results (Prevalence) |
| How was self-stigma conceptualised? |
| Impact of self-stigma (why-try effect, self-efficacy, self-esteem, treatment adherence, health needs, help-seeking) |
| Practical Recommendations |
| Future areas for research (gaps identified) |
| Additional relevant information |

